# ASSESSMENT OF POTENTIAL SARS-CoV-2 VIRUS N GENE INTEGRATION INTO HUMAN GENOME REVEALS NO SIGNIFICANT IMPACT ON RT-qPCR COVID-19 DIAGNOSTIC TESTING

**DOI:** 10.1101/2021.06.21.21258023

**Authors:** Erica Briggs, William Ward, Sol Rey, Dylan Law, Katharine Nelson, Michael Bois, Nili Ostrov, Henry H. Lee, Jon M. Laurent, Paolo Mita

**Author notes:** Corresponding author: Paolo Mita, Pandemic Response Lab, 30-02 48^th^ Avenue, Long Island City, Suite 360, New York, 11101, USA.

## Abstract

The SARS Coronavirus 2 (SARS-CoV-2) pandemic presents new scientific and scale-up challenges for diagnostic capabilities worldwide. The gold standard diagnostic for SARS-CoV-2 infection is a reverse transcription/quantitative PCR (RT-qPCR) which targets the viral genome, an assay that has now been performed on millions of patient specimens worldwide regardless of symptomatic status. Recently Zhang et al. suggested the possibility that the SARS-CoV-2 N gene could integrate into host cell DNA through the action of the LINE-1 retrotransposon, a mobile element that is potentially active in human somatic cells, thereby calling into question the veracity of N-gene based RT-qPCR for detection of SARS-CoV-2 infection. Accordingly, we assessed the potential impact of these purported integration events on nasal swab specimens tested at our clinical laboratory. Using an N-gene based RT-qPCR assay, we tested 768 arbitrarily selected specimens and identified 2 samples which resulted in a positive detection of viral sequence in the absence of reverse transcriptase, a necessary but not sufficient signal consistent with possible integration of the SARS-CoV-2 N gene into the host genome. Regardless of possible viral N gene integration into the genome, in this small subset of samples, all patients were still positive for SARS-CoV-2 infection, as indicated by a much lower Ct value for reactions performed in the presence of reverse transcriptase (RT) versus reactions performed without RT. Moreover, one of the two positives observed in the absence of RT also tested positive when using primers targeting ORF1ab, a gene closer to the 5’ end of the genome. These data are inconsistent with the N gene integration hypothesis suggested by the studies by Zhang et al., and importantly, our results suggest little to no practical impact of possible SARS-CoV-2 genome integration events on RT-qPCR testing.

**COMPETING INTEREST STATEMENT:** The authors of this study are employees of the Pandemic Response Lab (PRL)/ReOpen Diagnostics, a private company performing SARS-CoV-2 RT-qPCR based testing, an area of interest of this study.

## INTRODUCTION

SARS Coronavirus 2 (SARS-CoV-2) is an enveloped positive strand RNA virus that belongs to the beta Coronavirus family together with the Middle East Respiratory Syndrome coronavirus (MERS) and the Severe Acute Respiratory Syndrome coronavirus (SARS) [1]. Other members of the Coronavirus family that infect humans include HCoV-229E and HCoV-OC43 (alpha coronaviruses), HCoV-NL63 and HCoV-HKU1 (beta coronaviruses) and are the cause of mild respiratory tract infections associated with symptoms of the ‘common cold’ [2]. SARS-CoV-2 is the cause of coronavirus disease 2019 (COVID-19), a contagious respiratory illness characterized by a wide range of symptoms the severity of which ranges from complete asymptomatic disease course to acute respiratory pathologies requiring mechanical ventilation, and in some cases causing death. SARS-CoV-2 is transmitted through infected secretions such as saliva and respiratory secretions or respiratory droplets, which are expelled when an infected person coughs, sneezes, talks or sings (WHO Scientific Brief 9 July 2020) [3].

The SARS-CoV-2 virion is constituted by four structural proteins: Spike (S), Membrane (M), Nucleocapsid (N) and Envelope (E) proteins. SARS-CoV-2 enters the cells through the interaction of its spike protein with ACE2 (angiotensin I converting enzyme 2) and through cleavage-mediated processing of the spike protein by TMPRSS2 [4]. Other membrane proteins may mediate the virus interaction and entry into the target cell [5]. Upon binding to the attachment factors of the target cells, SARS-CoV-2 enters the cells inside a membrane vesicle/endosome. After uncoating, initial translation of the virus RNA is immediately initiated. The RNA genome of SARS-CoV-2 is more than 30 kb long and flanked by 5’ and 3’ untranslated regions (UTR) important for virus translation; two thirds of the RNA encodes 15-16 non-structural proteins (nsp) mainly involved in RNA replication and transcription and part of two big open reading frames (ORF1a and ORF1b) on the 5’ of the viral RNA. The 3’ one third of the viral genome encodes for structural proteins and accessory proteins. In the host cell cytoplasm, the replicated viral RNA genome and the translated structural proteins are assembled in new virion particles released from the cells by exocytosis through the Golgi [6].

Previous studies are in conflict about the potential for SARS-CoV-2 to integrate into the human genome. One recent study suggested that the LINE-1 (Long Interspersed Nuclear Element-1) retrotransposon may reverse transcribe the SARS-CoV-2 RNA genome, or part of it, into the genome of the host cell [7]. LINE-1 retrotransposons are genetic elements able to “copy and paste” themselves in the human genome through a process generally called “retrotransposition”. As a consequence of retrotransposon activity during evolution, 21% of our genome is made up of LINE-1 sequences [8, 9]. However, the vast majority of these sequences are inactive and no longer able to move. Only 100 copies of LINE-1 are full length and maintain the ability of moving to new genomic locations [10]. To counteract this potentially dangerous activity, mammalian host cells tightly and efficiently repress the expression of LINE-1 through a plethora of transcriptional and post-transcriptional mechanisms in somatic cells. Activity of LINE-1 has been recorded during early stages of development, in some specific cellular contexts such as oocyte attrition or neurogenesis, and under stress conditions such as aging and pathological conditions including cancer [11-15]. Recent experiments suggest activity of LINE-1 elements in epithelial cells [16]. *In vitro* experiments have shown that, in conditions of high LINE-1 de-repression and expression, the machinery that mediates LINE-1 retrotransposition, can mobilize other sequences such as *Alu* and SVA elements, and can even lead to integration of certain cellular mRNAs, leading to the formation of processed pseudogenes [17-20]. One study even showed that LINE-1 elements can promote the low frequency integration of certain RNA viruses [21]. It is therefore formally possible that, in cells infected by SARS-CoV-2, which might lead to compromised genome regulation allowing for LINE-1 de-repression, SARS-CoV-2 RNA molecules might be bound by the LINE-1 machinery and integrated into the host cell genome. This event was postulated to have occurred in autopsied lung samples from 14 COVID-19 patients, in 4 lung organoids preparations as identified by viral-human chimeric RNA measured by RNA-seq and in cell lines artificially over-expressing LINE-1, as identified by viral-human chimeric DNA measured by genome sequencing [7]. The chimeric (virus/human) reads from human samples used to support the hypothesis of viral integration into human genome were 0.004-0.14% of the total SARS-CoV-2 [7] reads, representing a low percentage of the total reads [22]. No significant chimeric reads were found in bronchoalveolar lavage samples, and nasal swabs, the usual mode of collection for PCR-based testing, were not analyzed in that study. These findings have potentially significant consequences in interpretation of RT-qPCR measurement of SARS-CoV-2 RNA in patient nasal samples, today’s gold standard procedure for the identification of SARS-CoV-2 infection and COVID-19 diagnosis [23]. The postulated LINE-1 mediated insertion of SARS-CoV-2 genome into the host DNA would particularly impact PCR-based tests utilizing primers/probes targeting the N gene at the 3’ end of SARS-CoV-2 genome because of the 3’ bias of LINE-1 insertions [24] and the high expression of the SARS-CoV-2 N gene [25, 26]. Indeed, all of the events observed by Zhang et al. involved the highly expressed N gene.

Other studies suggested different interpretations for the data identifying virus/host chimeric reads after next generation sequencing (NGS) of SARS-CoV-2 infected cells. These works pointed out the possibility of fusion events happening during library preparation [22, 27] that could explain the nature of the identified chimeric DNA fragments containing both human and viral sequences. Specifically, Yan et al. [22] found that the observed chimeric events [7] fell below background and were likely due to template switching of the reverse transcriptase enzyme used during RNA-seq library preparation. They concluded that the chimeric events observed were rare, unreproducible, and unlikely to have occurred biologically [22]. Moreover, a recent study showed no evidence of SARS-COV-2 integration into the genome of infected HEK293T cells using long-read Oxford Nanopore Technologies (ONT) sequencing [28].

Integration of SARS-CoV-2 genes into the human genome, if it occurs, may have significant consequences in the interpretation of standard procedure for identification of SARS-CoV-2 infection and COVID-19 diagnosis [23]. This is because the gold-standard RT-qPCR test, which uses both reverse transcriptase and DNA polymerase in a one-step reaction, is meant to measure expression of viral genes from viral RNA, and would not discriminate between that and expression of a viral gene that is integrated in the human genome. Thus, integration of viral genes would result in false positive test results.

To evaluate the practical relevance of the hypothesis put forward by Zhang et al. [7] on diagnostic testing for COVID-19, we searched for evidence of viral N gene integration into the host genome by analyzing 768 remnant, de-identified COVID positive nasal swab samples submitted to the Pandemic Response Lab (PRL) NYC for SARS-CoV-2 testing and sequencing. We performed a two step RT-PCR reaction in the presence or absence of reverse transcriptase (RT) enzyme. Our results show marginal (2/768 positive samples) evidence of SARS-CoV-2 DNA. These data suggests that, if it is happening at all, LINE-1 mediated retrotransposition of SARS-CoV-2 genome into the host DNA is such a rare and low frequency event with no practical impact on the RT-PCR-based diagnostic capability of the most widely used tests using the N gene primer probe sets targeting the sequences purportedly integrated into the genome according to Zhang et al [7].

## RESULTS

### Evaluation of SARS-CoV-2 sequences in host DNA

PCR-based testing of SCOV2 implements a one-step reaction that combines the reverse transcriptase (RT) and DNA amplification reaction in a single well, and therefore cannot distinguish between amplification of RNA or DNA targets. According to Zhang et al. [7], the LINE-1 reverse transcriptase mediated integration of SARS-CoV-2 DNA is almost exclusively limited to the 3’ region of the genome, and specifically of the N gene, as this is the most highly abundant viral mRNA found in infected human cells [25, 26]. For this reason, we compared the performance of primer sets either targeting this region (N1, N2 and N4 primer sets) or a 5’ SARS-CoV-2 region that should not be affected by the postulated insertion (ORF1ab) [29]. Patient samples containing cells that underwent SARS-CoV-2 integration into their genome, should be easily distinguishable from those that have not, since amplification of target viral sequences will occur in the absence of the reverse transcriptase (RT) step of COVID-19 RT-PCR diagnostic testing.

To assess the potential for SARS-CoV-2 integration into the host genome, we conducted two step RT qPCR, composed of an RT step followed by DNA amplification and quantification by qPCR. We used nucleic acids extracted from 192 positive SARS-CoV-2 patient samples collected from nasal swabs and tested by RT PCR using N1 and N2 primer probe sets on the PRL production line in Manhattan, New York. After completion and reporting of test results, multiwell plates containing the residual extracted RNAs from both negative (∼97-98.7% of samples) and positive (∼1.3-3.0% of sample) patients were transported on dry ice to PRL’s Long Island City, R&D labs for evaluation. Positive samples from multiwell plates were consolidated into 384 well plates using Opentron OT2 liquid handlers. The consolidated positive samples had a broad bimodal Ct distribution, representative of large scale testing at PRL (Supplemental Figure S1). The analyzed samples were also part of PRL SARS-CoV-2 sequencing effort; the distribution of the SARS-CoV-2 variants is presented in Figure S2. After consolidation of the positive samples into 384 well plates, samples were tested using both N1 and N2 primer sets with and without reverse transcriptase enzyme, and then evaluated for amplification by probe-based qPCR (Figure 1A and B). We expect samples treated with reverse transcriptase to be able to amplify SARS-CoV-2 RNA and DNA during qPCR, while samples without reverse transcriptase would only be able to amplify DNA present in the reaction. In this initial evaluation experiment, we found 155 of the 192 samples produced positive COVID19 results, representing 80.7% of the initial diagnostic one-step RT-qPCR, demonstrating reduced efficiency and sensitivity when moving from our optimized one-step reaction to the two-step procedure. Additional factors may also include sample freeze-thaw cycles and the use of only one-fifth the RNA input volume, due to limited remnant samples, compared to our clinical testing pipeline. Following the 2-step RT-PCR we identified four samples that amplified N1 or N2 sequences in the absence of RT (Table 1). While this initial result suggested the possibility of SARS-CoV-2 integration, we could not eliminate the possibility of contamination with RT enzyme, SARS-CoV-2 amplicons, not unlikely in research settings compared to the higher standards of diagnostic labs (see discussion), or to the intrinsic but low level RT activity of Taq polymerase in the absence of RT [30].

**Figure 1.**
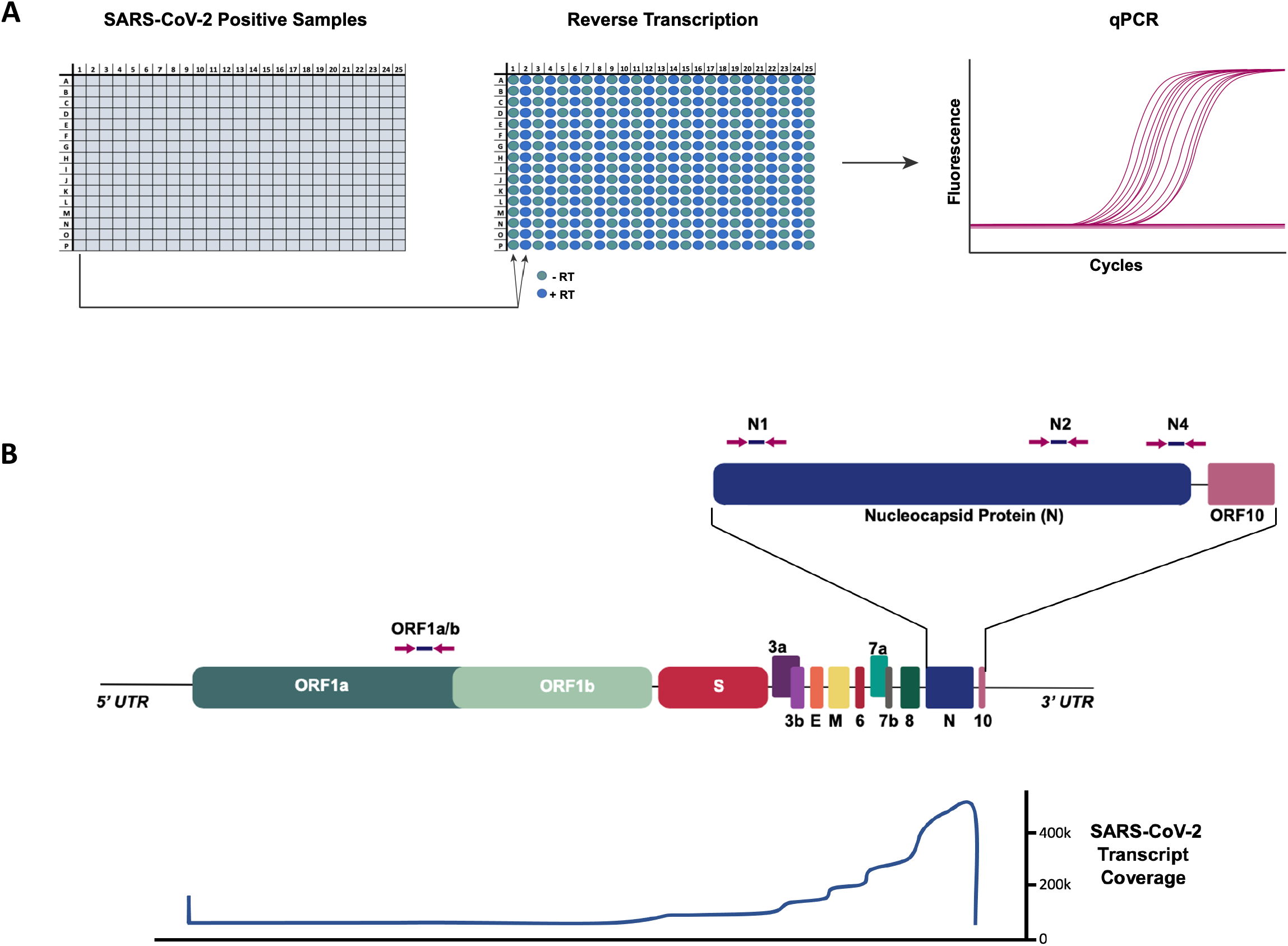
Analysis of SARS-CoV-2 positive samples for genomic SARS-CoV-2 DNA. **A)** SARS-CoV-2 positive samples were consolidated in a 384 well plate. Each sample was run in duplicate reverse transcription (RT) reactions with or without reverse transcriptase. Resulting reactions were assessed for SARS-CoV-2 cDNA by qPCR. **B)** (top)Schematic of SARS-CoV-2 genome. qPCR primer/probe locations for N1, N2, N4 and ORF1ab are shown with arrows. (bottom) Profile of RNA read depth across the viral genome as reported by [25, 26]

**Table 1.** Initial RT-qPCR evaluation for SARS-CoV-2 DNA. RT-qPCR was performed on SARS-CoV-2 positive samples either with (+RT) or without (-RT) reverse transcriptase (as described in Figure 1). qPCR for N1 and N2 were performed and numbers of positive samples and average Ct values (for -RT positive reaction and for total reactions) are shown (raw data are presented in Supplemental Table 2).

|  | **Number of Samples**<br>**(Percent of +RT)** | **Average Ct**<br>**(-RT Amplified Samples)** | **Average Ct**<br>**(Total Samples)** |
| --- | --- | --- | --- |
| **N1 +RT** | 139 | 21.09 | 24.54 |
| **N1 -RT** | 4 (2.88%) | 32.28 | 32.38 |
| **N2 +RT** | 112 | 20.40 | 24.24 |
| **N2 -RT** | 2 (1.79%) | 29.26 | 29.26 |
| **RP +RT** | 187 | 23.06 | 24.99 |
| **RP -RT** | 188 | 27.99 | 29.54 |
| **Samples Tested** | 192 |  |  |

SARS-CoV-2 integration was previously attributed to the activity of the LINE-1 retrotransposon which purportedly selectively integrates the N gene region only. Consistent with the mechanism of LINE-1 retrotransposition, as well as the high expression level of the N gene, Zhang et al. described a higher frequency of SARS-CoV-2 chimeric transcripts that included the 3’ end of the SARS-CoV-2 genome and more specifically the N gene. To further evaluate positive SARS-CoV-2 samples for genomic insertions, we tested an additional 576 SARS-CoV-2 positive nasal samples with and without reverse transcriptase (Table 2A). We also included two additional primer/probe targets, N4 and ORF1ab, to the follow-up analyses. The N4 primers/probe partially target the nucleocapsid protein, but, compared to N1 and N2 primers/probes, they are closer to the 3’ end of SARS-CoV-2 genome. N4 is therefore more likely to amplify samples with viral genomic integration (Figure 1B). ORF1ab primers/probe targets a region closer to the 5’ end of the SARS-CoV-2 genome, a region where Zhang et al observed little or no evidence of SARS-CoV-2 integration into the host genome. In total, we observed 22 samples (4 from experiment in Table 1 and 18 from experiment in Table 2A) with -RT amplification of at least one of the SARS-CoV-2 primers (N1, N2, N4, or ORF1ab) (Table 2B). Of these 22, only six samples amplified the N4 target, four of which also included N1 or N2 amplification. We consistently observed a much higher Ct value for the samples amplified in the absence of RT reactions compared to their +RT counterparts (Tables 1 and 2), suggesting the presence of abundant viral RNA, even when SARS-CoV-2 DNA was possibly present. This indicates that all patients reported as positive for Coronavirus RNA were infected with SARS-CoV-2 virus during the time of sample collection, regardless of possible genomic integration of SARS-CoV-2. This result is in line with Zhang et al., who observed SARS-CoV-2 N gene insertion in samples from patients with active SARS-CoV-2 infection [7]. To further explore samples that had amplification in the absence of RT, we repeated the minus RT reaction using the remaining extracted RNA/DNA and found that only two reactions repeated (.26% of total positive samples). We also ran the reaction in the presence or absence of DNase to test if any possible amplification was due to the presence of target DNA. As expected, treatment with DNase eliminated the observed signal in the absence of RT condition as well as the positive RPP30 gene, a standard control for human DNA used as internal control. Despite the fact that these data and experiments do not address whether genomic integration of SARS-CoV-2 N gene is possible, it suggests a marginal impact, if any, on SARS-CoV-2 RT-qPCR testing from nasal swab samples.

**Table 2.**
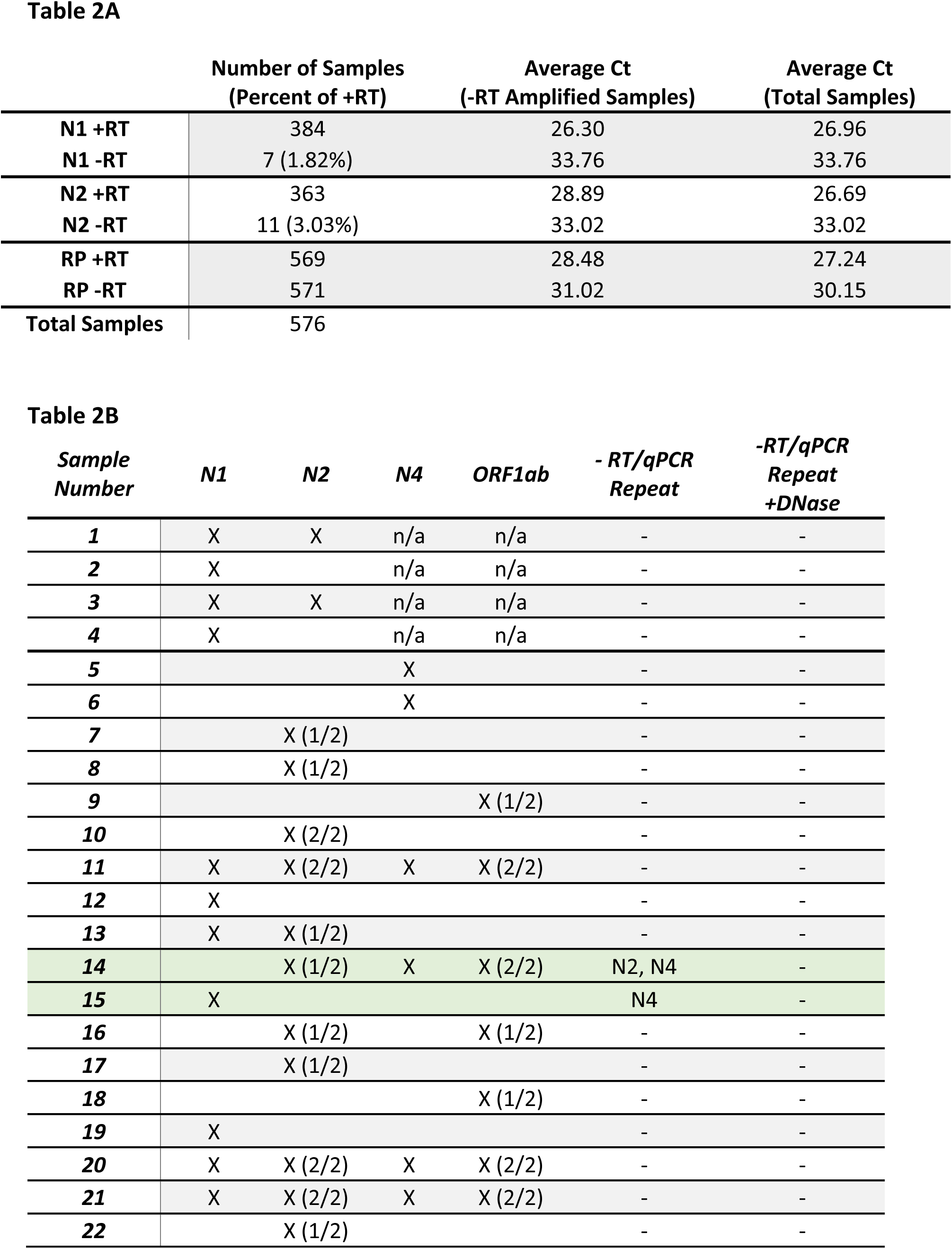
Comprehensive RT-qPCR evaluation for SARS-CoV-2 DNA. **A)** RT-qPCR was performed on SARS-CoV-2 positive samples either with (+RT) or without (-RT) reverse transcriptase (as described in Figure 1). qPCR for N1, N2, N4 and ORF1ab was performed and number of positive samples and average Ct values are shown. **B)** Comprehensive table of all SARS-CoV-2 amplifications in the -RT condition, including our initial evaluation (Samples 1-4) and our comprehensive evaluation (Samples 5-22). Parentheses for N2 and ORF1ab display repetition in technical qPCR replicates. -RT/qPCR Repeat and -RT/qPCR Repeat +DNase columns include results for a repetition of the reverse transcriptase reaction in the absence of RT and qPCR for N1, N2, N4 and ORF1ab (- indicate no amplification detected) (all raw data are presented in Supplemental Table 2 and summary analysis in Supplemental Table 3).

## MATERIAL AND METHODS

### Reverse Transcription quantitative PCR (RT-qPCR)

#### SARS-CoV-2 RT-PCR test

The *PRL SCV2 test* is a real-time reverse transcription polymerase chain reaction (rRT-PCR) clinical test. The SARS-CoV-2 primer and probe set is designed to amplify and detect RNA from the SARS-CoV-2 in nasopharyngeal, nasal, oropharyngeal swabs and saliva specimens from patients as recommended for testing by public health authority guidelines. The assay employs the CDC-designed primer/probe sequences as included in the SARS-CoV-2 (2019-nCoV) CDC qPCR Probe Assay in Supplemental Table 1 (IDT, 2019-nCoV CDC Primers and Probes). Two separate regions of the viral nucleocapsid (N) gene are targeted by the PRL SCV2 test (N1 and N2). Also included is an internal control targeting the human RNase P/RPP30 (RP) gene. All three targets are detected in a single assay in multiplex, each with a unique fluorophore-quencher combination as outlined in Supplemental Table 1.

Nucleic acids (DNA/RNA) are isolated and the SARS-CoV-2 RNA is then reverse transcribed and amplified together with the RNA P control using a one-step RT-qPCR. Taq-man real-time RT-PCR is performed. Five microliters of extracted RNA are used in a 10µl one step-RT-qPCR reaction. Positive specimens for SARS-CoV-2 RNA have a Ct value lower than 37 for the N1 and/or N2 primer/probe set. Curves with an Amplification Ration (AR= (fluorescence at cycle 50)/(fluorescence at cycle 5)) smaller than 1.5 were considered negative.

### Two step RT-PCR

All experiments were performed using remnant RNA/DNA samples previously confirmed by PRL SARS-CoV-2 test to be COVID-19 positive. Patients’ samples were consolidated in 384 well plates using an OT2 liquid handler (Opentrons). Five microliters of each sample were used for each of 2 RT reactions: (I) reaction without Reverse Transcriptase enzyme, (II) reaction with Reverse Transcriptase enzyme (Takara PrimeScript RT reagent kit # RR037B and Takara Pre-mix Ex Taq master mix for probe-based real-time PCR # RR390W). Two microliters of RT reaction were used in a 10µl qPCR-PCR reaction using the CDC N1 and N2 primers and probes as well as additional primers and probes targeting ORF1ab (CDC China set, [29]) and N4 set (CDC’s Influenza SARS-CoV-2 Multiplex Assay, SC2 CDC FluViD assay, IDT) (Supplemental Table 1).

For DNase treatments before RT reactions the Primescript RT Reagent Kit with gDNA Eraser (Perfect Real Time) (Takara #RR047A) was used.

### Genome Assembly (PRL)

For each specimen, sequencing adapters are first trimmed using Trim Galore v0.6.6 [31], then aligned to the SARS-CoV-2 Wuhan-Hu-1 reference genome (NCBI Nucleotide NC_045512.2) using BWA MEM 0.7.17-r1188 [32]. Reads that are unmapped or those that have secondary alignments are discarded from the alignment. Consensus and mutations were called using samtools and Intrahost variant analysis of replicates (iVar) [33] with a minimum quality score of 20, frequency threshold of 0.6 and a minimum read depth of 10x coverage. A consensus genome with ≥ 90% breath-of-coverage with ≤ 3000 ambiguous bases is considered a successful reconstruction (as per APHL recommendation). Variants were called using PANGOLIN [34] v2.1.11 to v2.3.8.

### Library preparation and sequencing (PRL)

Positive RNA specimens between cycle threshold of 15-30 were selected from all samples tested at Pandemic Response Labs, NYC and cDNA for each specimen was generated using LunaScript RT SuperMix (NEB, MA) according to manufacturer protocol. To target SARS-CoV-2 specifically, cDNA for each specimen was amplified in two separate pools, 28- and 30-plex respectively, to generate 1200bp of overlapping amplicons [35] using Q5 2x Hot-Start Master Mix (NEB, MA). The resulting pools are combined in equal volume and enriched for full length 1200 bp product using a SPRI-based magnetic bead cleanup. Enriched amplicons are tagmented (Illumina, CA) and barcoded (IDT, IA) and paired-end sequenced on an Illumina MiSeq or NextSeq 550.

## DISCUSSION

A recent study suggested the possibility of SARS-CoV-2 N gene mRNA reverse transcription and integration into host cell DNA by LINE-1 retrotransposon [7]. Apart from data obtained from cell lines (HEK293T and Calu3 cell lines), an *in vitro* and artificial setting not necessarily recapitulated in an *in vivo* context, viral/human chimeric reads were identified after analysis of RNA sequencing of 14 autopsied lung samples and 4 lung organoid preparations from COVID 19 patients. These reads represented 0.004-0.14% of the total SARS-COV-2 and, consequently, an even lower percentage of the total genomic DNA reads [7]. Follow-up studies have questioned whether SARS-CoV-2 integration into the human genome actually occurs at all. In depth sequencing analysis suggests that most if not all mRNA/human nuclear DNA chimeric reads are due to artifacts generated during RNA-seq library prep [22], while Zhang et al. argue that SARS-CoV-2 chimeric transcripts are genuine and present in patient samples [7]. Also, deep (>50×) long-read Oxford Nanopore Technologies (ONT) sequencing of HEK293T cells genomes infected with SARS-CoV-2, found no evidence of viral insertion into the genome [28]. Since the predominant diagnostic test for SARS-CoV-2 is reverse transcription quantitative PCR (RT-qPCR) using N gene probe sets (N1, N2 from USA CDC SARS-CoV-2 test), the presence of N gene sequences in the human genome would have the potential to produce false positive results after a patient recovers from an actual infection. Evaluating this possibility and its impact on RT-qPCR testing is vital for maintaining accuracy in SARS-CoV-2 diagnostic testing.

We assessed the potential for the detection of N gene integration using a standard PCR based test by performing a two-step RT-qPCR with and without reverse transcriptase enzyme. While the two-step reaction differed slightly from the diagnostic one step RT-qPCR (i.e., higher mean Ct, and somewhat less sensitive), we initially identified 22 of 768 samples showing SARS-CoV-2 amplification in the absence of reverse transcriptase. However, when repeated, only two of these samples, Sample 14 and Sample 15, continued to have amplification without RT and, of those two, only one in an N gene specific way. Sample 14 showed an amplification signal for N2, N4 and ORF1ab. According to the findings of Zhang et al, the insertion of ORF1ab was highly unlikely, suggesting this is unlikely to represent a SARS-CoV-2 integration. The second sample, Sample 15, amplified N1 and N4 in the absence of reverse transcriptase, potentially supporting the hypothesis for N mRNA integration. While we cannot eliminate the possibility of N mRNA integration in these two samples, these patients also had high levels of SARS-CoV-2 RNA as evidenced by the much lower +RT Ct values (Table 2A) indicating it was correctly reported as SARS-CoV-2 positive. Additionally, other potential sources of amplification may be producing false positives in our -RT condition. These samples were processed downstream of the PRL diagnostic test in PRL’s non-diagnostic research and development facility, a laboratory that routinely preforms SARS-CoV2 PCR, creating the potential for SARS-CoV-2 amplicon contamination. Amplicon contamination is indeed a likely event in research laboratories performing high numbers of PCR against specific targets [36]. At PRL’s diagnostic lab in Manhattan, many quality controls and periodic checks, as well as strict behavioral and methodological procedures, are implemented regularly to limit, identify and resolve possible false positive sources. This standard of quality cannot be maintained in most if not all research labs that should always consider the rate of amplicon contamination when performing high numbers of RT-qPCR. Additionally, Taq polymerase has been shown to reverse transcribe RNA in specific buffers, potentially giving rise to false positives in our -RT condition [30]. However, this is a quite unlikely hypothesis considering that the RT-independent/Taq-dependent signal would be expected to be present in all reactions.

Overall, our results show minimal evidence, if any, of N gene integration and an insignificant impact on RT-qPCR based COVID testing from nasal swab samples.

## Data Availability

All relevant data are submitted with this manuscript

## FIGURE LEGENDS

**Supplemental Table 1.**
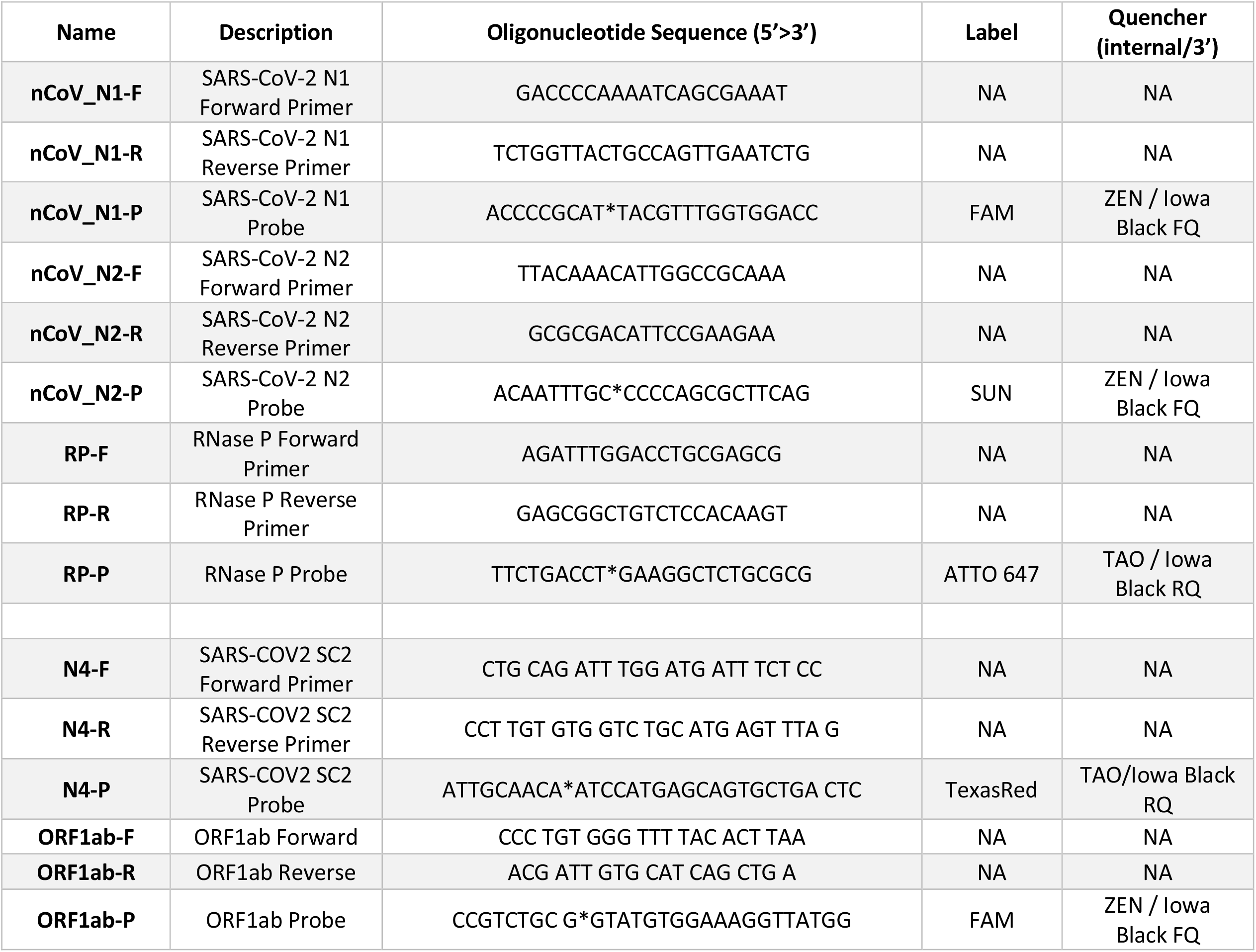
Primer and probe sequences with corresponding fluorophores and quenchers used in this study are reported.

**Supplemental Table 2.** (Expansion of Table 2B) Comprehensive table of all SARS-CoV-2 amplifications in the -RT condition, including our initial evaluation (Samples 1-4) and our comprehensive evaluation (Samples 5-22). Includes Ct values for +/- RT qPCR, variant status as determined by sequencing, and results of DNase treatment.

| Sample Number | N1 | N2 | N4 | ORF1ab | N1 Ct (+RT) | N1 Ct (-RT) | N2 Ct (+RT) | N2 Ct (-RT) | N4 Ct (+RT) | N4 Ct (-RT) | ORF1ab Ct (+RT) | ORF1ab Ct (-RT) | Variant | -RT qPCR Repeat | -RT qPCR +DNase Repeat |
| --- | --- | --- | --- | --- | --- | --- | --- | --- | --- | --- | --- | --- | --- | --- | --- |
| 1 | X | X | n/a | n/a | 18.56 | 30.68 | 19.11 | 31.61 |  |  |  |  | B.1.1.7 | - | - |
| 2 | X |  | n/a | n/a | - | 35.80 | - | - |  |  |  |  |  | - | - |
| 3 | X | X | n/a | n/a | 21.13 | 26.55 | 21.68 | 26.91 |  |  |  |  | B.1.526 | - | - |
| 4 | X |  | n/a | n/a | 23.57 | 36.50 | 23.64 | - |  |  |  |  | B.1.526.2 | - | - |
| 5 |  |  | X |  | 28.56 | - | 28.26 | - | 30.01 | 37.47 | 28.51 | - |  | - | - |
| 6 |  |  | X |  | 24.21 | - | 23.83 | - | 25.52 | 37.55 | 25.05 |  |  | - | - |
| 7 |  | X (1/2) |  |  | - | - | 34.11 | 36.40 | 34.23 | - | - | - |  | - | - |
| 8 |  | X (1/2) |  |  | - | - | 33.91 | 35.30 | - | - | - | - |  | - | - |
| 9 |  |  |  | X (1/2) | - | - | - | - | - | - | - | 32.06 |  | - | - |
| 10 |  | X (2/2) |  |  | - | - | 33.46 | 35.55 | - | - | - | - |  | - | - |
| 11 | X | X (2/2) | X | X (2/2) | 24.17 | 28.31 | 27.88 | 28.58 | 26.64 | 30.86 | 28.50 | 29.24 |  | - | - |
| 12 | X |  |  |  | 33.87 | 41.40 | - | - | - | - | - | - |  | - | - |
| 13 | X | X (1/2) |  |  | 26.20 | 36.80 | 26.54 | 37.35 | 28.68 | - | 26.95 | - |  | - | - |
| 14 |  | X (1/2) | X | X (2/2) | 32.76 | - | 32.24 | 36.00 | 33.05 | 36.09 | - | 34.30 |  | N2, N4 | - |
| 15 | X |  |  |  | - | 35.33 | - | - | - | - | - | - |  | N4 | - |
| 16 |  | X (1/2) |  | X (1/2) | 33.14 | - | 32.79 | 36.97 | 32.48 | - | - | 34.05 |  | - | - |
| 17 |  | X (1/2) |  |  | - | - | - | 33.50 | - | - | - | - | P.1 | - | - |
| 18 |  |  |  | X (1/2) | 31.95 | - | 31.10 | - | 33.34 | - | - | 32.74 |  | - | - |
| 19 | X |  |  |  | 31.56 | 36.62 | 45.00 | - | - | - | - | - |  | - | - |
| 20 | X | X (2/2) | X | X (2/2) | 21.13 | 31.64 | 21.90 | 33.52 | 23.48 | 34.22 | 22.01 | 31.85 | B.1.617.2 | - | - |
| 21 | X | X (2/2) | X | X (2/2) | 20.87 | 31.82 | 21.36 | 32.24 | 23.07 | 33.84 | 21.76 | 32.00 | P.1 | - | - |
| 22 |  | X (1/2) |  |  | 24.46 | - | 24.67 | 38.55 | 27.29 | - | 26.63 | - | B.1.1.7 | - | - |

**Supplemental Table 3.**
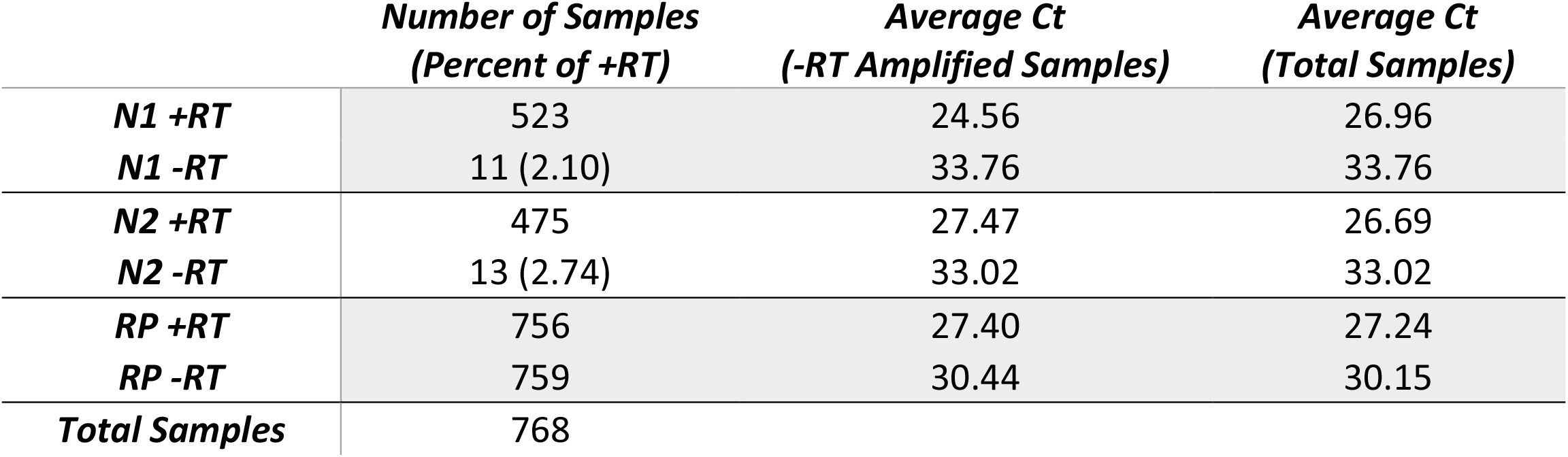
Combined summary analysis of initial and comprehensive RT-qPCR analyses in the presence (+RT) or absence (-RT) of reverse transcriptase.

**Supplemental Figure 1.**
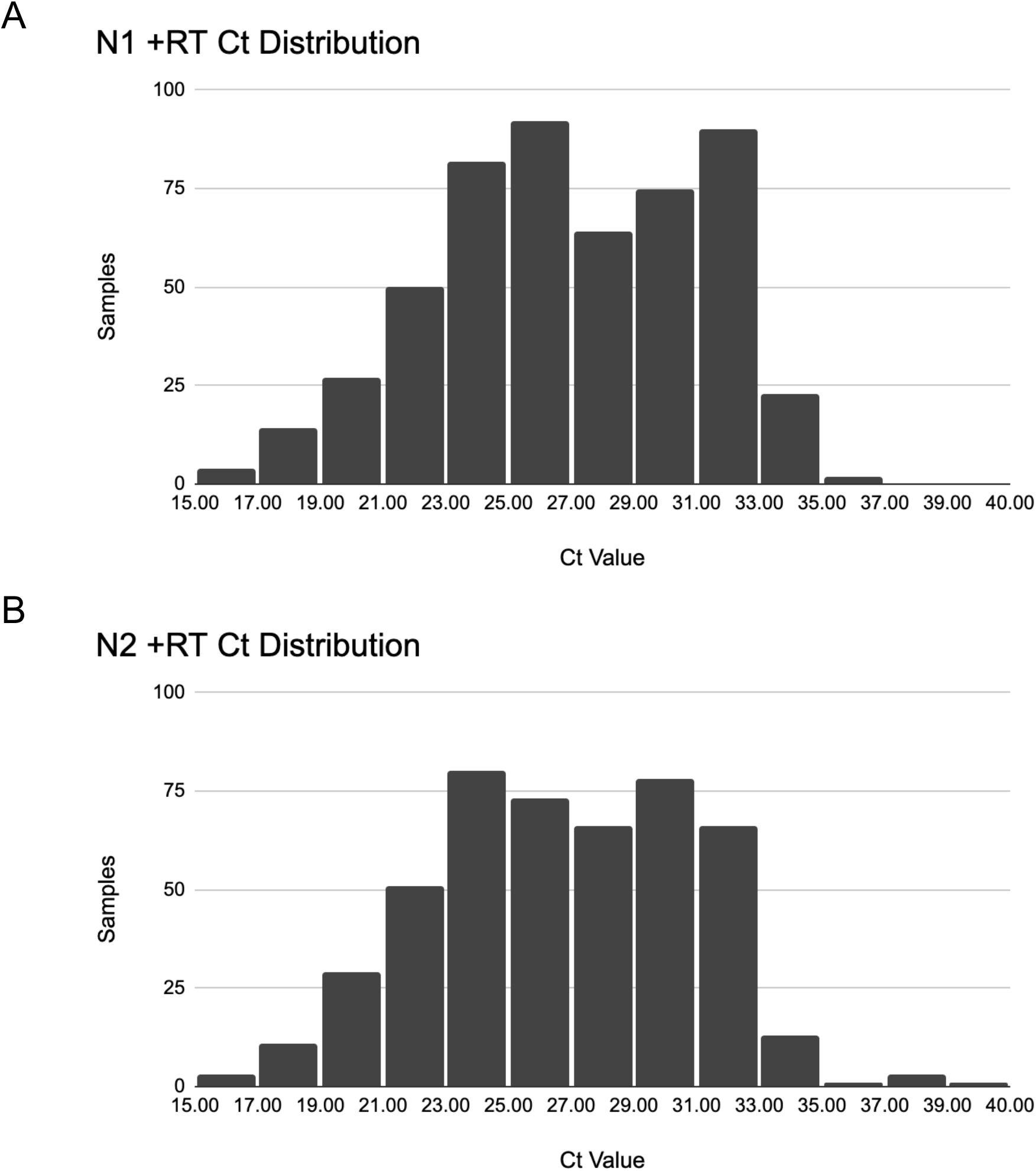
Ct distribution of positive SARS-CoV-2 samples used in this study in the presence of reverse transcriptase (+RT) for N1 (A) and N2 (B) as for PRL-SCV2 initial one-step RT-PCR test.

**Supplemental Figure 2.**
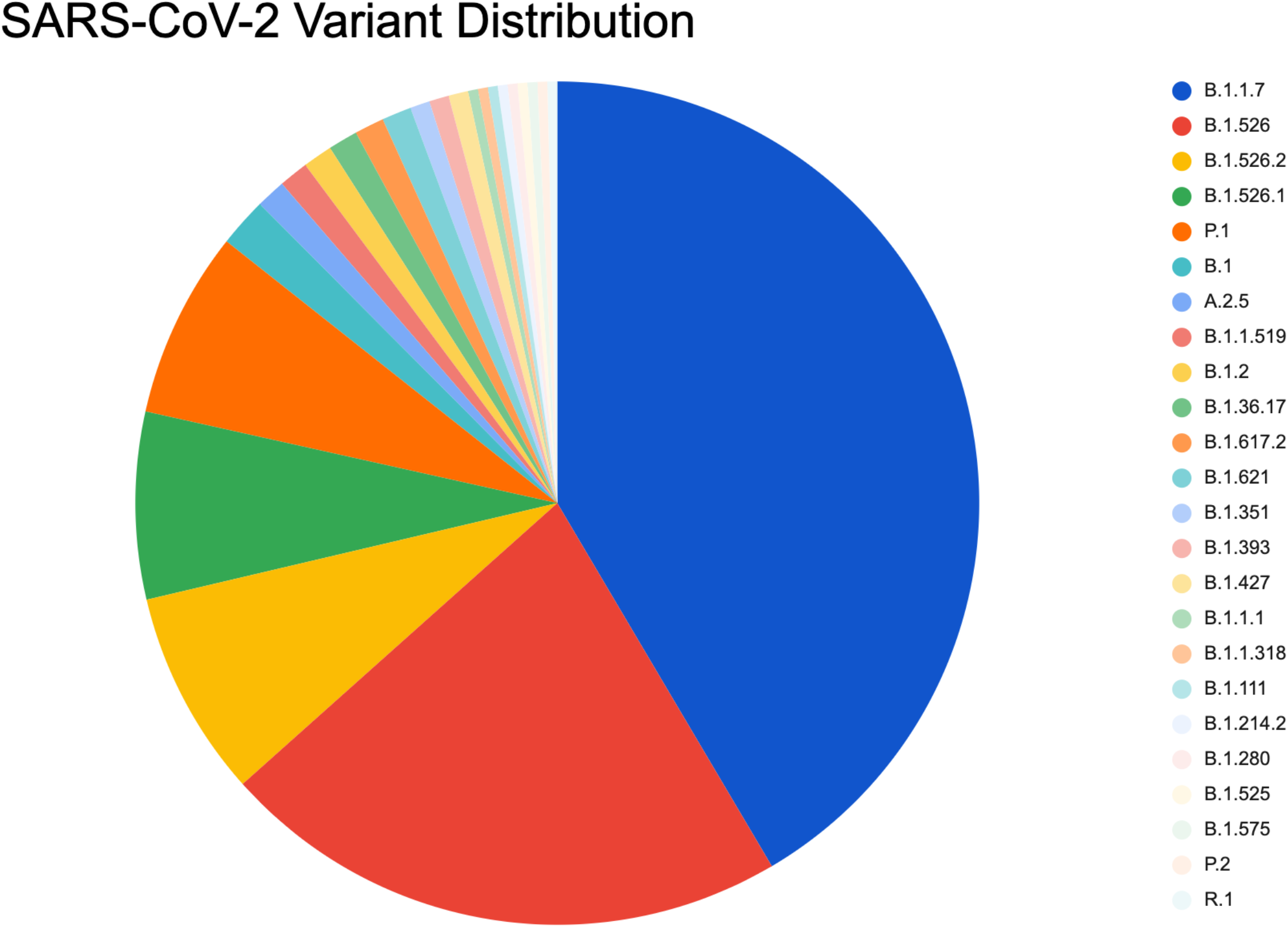
Distribution of SARS-CoV-2 variants among the samples analyzed in this study that underwent successful genome reconstruction of the virus genome, as [37].

